# Seasonal trends in colchicine prescriptions in England 2014-2019

**DOI:** 10.1101/2020.06.10.20127696

**Authors:** Mark Gibson

## Abstract

**Objectives:** To examine seasonal trends in the prescription of colchicine

**Methods:** A retrospective cohort study using a population level dataset of all community prescriptions for colchicine dispensed in England between December 2014 and November 2019.

**Results:** Significant seasonal variation exists in colchicine prescriptions (p<0.0001). Colchicine prescriptions were maximal in the summer months (June, July) in each of the years studied and lowest in the winter. Significant variation in colchicine prescriptions was observed between the summer and the winter(p<0.001), spring (p=0.0177) and autumn(p<0.001) months.

**Conclusion:** Seasonal trends in colchicine prescribing closely mirror previously observed seasonal trends in acute gout incidence providing further evidence that acute gout demonstrates seasonality and, in England, is more common in the summer.

## Introduction

Despite the availability of highly effective treatment gout, remains the most common form of inflammatory arthritis worldwide, although incidence varies by geographic region[1]. In the England acute gout now accounts for more hospital admissions than rheumatoid arthritis[2].

Seasonal variation in acute gout attacks has been observed but studies have shown seasonality differs by geographic location. A 2009 study of the Royal College of General Practitioners Weekly Returns Service (WRS), encompassing data from approximately 100 primary care practices across England and Wales, found that acute gout attacks peaked in the summer period of each year[3]. This observation was replicated in data from Korea[4], but contrasts with other studies showing acute gout was more common in the spring in Italy[5] and in autumn in Australia[6]. Studies from the United States of America are conflicting, with some demonstrating peak incidence in the autumn[7] and others in the spring[5,8].

This study aimed to explore seasonality in prescriptions for medication used to treat acute gout attacks in England. National guidelines produced by the British Society of Rheumatology outline treatment recommendations for the management of acute gout attacks and include the option of using strong non-steroidal anti-inflammatory drugs, colchicine or corticosteroids[9]. These recommendations are echoed in international guidance from both the European League against Rheumatism[10] and the American College of Rheumatology[11]. Of the recommended medications used to treat acute gout attacks Colchicine was chosen for the purposes of this study due to its higher specificity.

## Method

### Study design

This retrospective cohort study of English NHS primary care prescribing data analysed monthly prescription data for the 5-year period from December 2014 through to November 2019, the latest data available at the time of analysis. In keeping with internationally accepted practice, no ethical approval was required for this study as the data used is open, anonymised, and publicly available.

### Data sources

Monthly prescription data is made publicly available by the NHS England Business Services Authority. This dataset includes all primary care prescriptions that are dispensed in pharmacies in England. A normalised version of this data is provided by openprescribing.net[12], a freely available online resource. Data for all colchicine prescriptions between December 2015 and November 2019 was extracted from the openprescribing.net database by selecting BNF codes starting with 1001040G0.

Prescription data was adjusted to reflect the population size of the United Kingdom using mid-year estimates provided by the Office for National Statistics (2014-2018) [13]. The mid-year population estimate for the year 2019 was estimated assuming similar rates of population increase observed in both 2017 and 2018 (0.64%). The number of prescriptions of colchicine is reported as total items dispensed per 100,000 population. Prescription data for NSAIDs and corticosteroids were not included in this study due to the high number of indications other than acute gout.

### Statistical analysis

Seasonality was first assessed using the χ2-Goodness-of-Fit test on aggregated monthly prescription data. The null hypothesis assumed a uniform distribution with no variation between monthly prescriptions. Seasonal indices were computed for monthly colchicine prescription rates per 100,000 population. Monthly data was aggregated into seasons as follows: Winter (December-February), Spring (March-May), Summer (June-August), and Autumn (September-November). Statistically significant variation between seasons was assessed using one-way ANOVA and Tukey’s Range Test. A p-value of less than 0.05 was considered statistically significant.

Python 3 was used for data management (Pandas, Matplotlib, NumPy) and statistical analyses (SciPy).

## Results

Between December 2014 and November 2019 colchicine prescription rates in England showed statistically significant variation by χ2-Goodness-of-Fit test (p=0.0248). In each of the 5 years studied colchicine prescriptions were maximal in either June or July (Figure 1). Seasonality indices revealed colchicine prescriptions increased in the period May-August and decreased during the period November-February with prescriptions peaking in July (Figure 2). Statistically significant variation in colchicine prescription rates was observed between seasons (ANOVA, p<0.0001). Tukey’s Range Test revealed statistically significant differences were observed between summer-winter (p<0.001), summer-spring (p=0.0177), and summer-autumn (p<0.001). Statistically significant seasonal variation was also observed between autumn-spring (p=0.0156) and spring-winter (p<0.001). No significant variation was observed between autumn-winter (p=0.6132).

**Figure 1:**
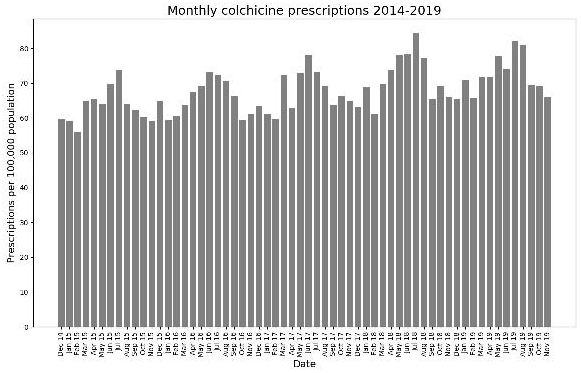
Trends in colchicine prescriptions dispensed in the community in England between December 2014 and November 2019. Prescription rates are given per 100,000 population.

**Figure 2:**
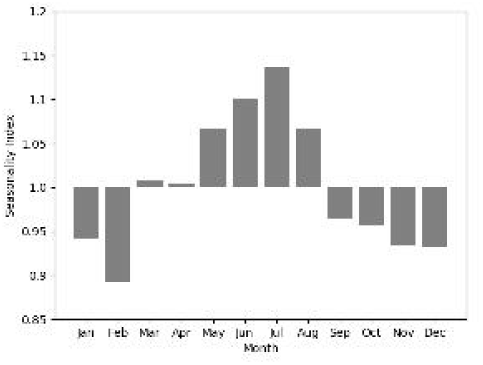
Seasonal indices calculated on monthly aggregated prescription data between December 2014 and November 2019

## Discussion

### Comparisons with other studies

The findings of this study closely mirror previous work demonstrating that in England acute gout was more likely in summer months and least likely in the winter[3]. The similar seasonal variation between both acute gout incidence and colchicine prescription rates provides validity for the use of colchicine prescriptions as a surrogate marker for the burden of acute gout attacks. This study provides further evidence of seasonal variation in acute gout.

### Findings in Context

The mechanism underlying the seasonal variation in acute gout is not well understood but several explanations have been proposed as to why acute gout may be more likely in the summer months. A study on seasonal variation in serum uric acid levels in a Swedish population revealed levels were highest in the period June-August[14]. Positive association between average ambient temperatures and risk of gout flare has been described[15] and it has been suggested that dehydration may precipitate intra-articular crystal formation[3].

In contrast with our findings, others have proposed explanations for why acute gout could be more common in colder months. *In vitro*, precipitation of uric acid crystals increases in colder temperatures[16]. It has been suggested that the dietary intake of purine rich foods increases in December as part of Christmas celebrations[7].

### Strengths and limitations

The primary strength of this study is the use of a large population-level dataset rather than sampled data. By analysing a record of every primary care prescription dispensed in England over the 5 year study period the seasonal trends of colchicine prescriptions in England can be clearly observed. The barriers associated with using NHS England’s open data have been discussed in detail recently[17]. Colchicine was chosen for use in this study because of its high specificity as a treatment for acute gout when compared to NSAIDs and corticosteroids but colchicine is not used exclusively for gout, with indications including the periodic fever syndromes and Behcet’s syndrome, potentially leading to confounding. The 2017 BSR guidelines for the management of gout advocate the use of colchicine or NSAIDs as prophylaxis when initiating urate lowering therapy potentially confounding the use of colchicine as a surrogate for acute gout. Hospital prescription data could not be included in this study as it is not currently published by the NHS England Business Services Authority.

### Future implications

In view of the increasing burden that acute gout attacks place on both primary and secondary care providers, further research into the factors that influence the seasonal variation in acute gout attacks is warranted. Identification of these factors may allow healthcare providers to better identify and manage those at risk of acute gout. Until such time where the factors behind the increased rates of acute gout in the summer months are understood gout sufferers in England should consider taking steps to remain well hydrated between the months of May and August.

### Key messages

□ There is seasonal variation in the prescribing of colchicine in England with prescriptions peaking in summer months
□ Seasonal trends in colchicine prescribing closely mirror previously observed seasonal trends in acute gout incidence, providing further evidence that acute gout demonstrates seasonality

## Data Availability

Monthly prescription data is made publicly available by the NHS England Business Services Authority. This dataset includes all primary care prescriptions that are dispensed in pharmacies in England. A normalised version of this data is provided by openprescribing.net, a freely available online resource.

https://openprescribing.net/

## Notes

### Competing Interest Statement

The authors have declared no competing interest.

### Funding Statement

This research did not receive any specific grant from funding agencies in the public, commercial, or not-for-profit sectors.

### Author Declarations

In keeping with internationally accepted practice, no ethical approval was required for this study as the data used is open, anonymised, and publicly available.

